# SARS-CoV-2 antibody response among COVID-19 patients is not affected by parasite co-infection

**DOI:** 10.1101/2023.08.20.23294345

**Authors:** Teklay Gebrecherkos, Yazezew Kebede Kiros, Feyissa Challa, Atsbeha Gebreegzabher, Saro Abdella, Dereje Leta, Abraham Desta, Ataklti Hailu, Geremew Tasew, Mahmud Abdulkader, Masresha Tessema, Getachew Tollera, Zekarias Gessesse Arefaine, Henk HDF Schallig, Britta C. Urban, Tobias F. Rinke de Wit, Dawit Wolday

## Abstract

**Background:** B-cell hypo-responsiveness has been associated with intestinal parasitic co-infection. The effect of parasite co-infection on antibody response to SARS-CoV-2 is unknown. Here, we aimed to determine antibody response to SARS-CoV-2 among COVID-19 patients co-infected with intestinal parasites and those without parasite co-infection.

**Methods:** In this prospective cohort study, a total of 589 samples were serially collected from 72 RT-PCR-confirmed patients. Anti-SARS-CoV-2 nucleocapsid protein (NP) antibody titers were measured longitudinally during hospitalization. SARS-CoV-2 infection was confirmed by RT-PCR on samples obtained from nasopharyngeal swabs, while direct microscopic examination, modified Ritchie concentration, and Kato-Katz methods were used to identify parasites and ova from fresh stool samples. Data were analyzed using STATA version 14.

**Results:** Of the 72 COVID-19 patients, 39 (54.2%) were co-infected with intestinal parasites while 33 (45.8%) had no parasitic co-infection. Overall, the median cut-off index (COI) for anti-NP antibody titer among COVID-19 patients co-infected with parasites was 6.91 (IQR: 0.55-40.7) compared to 7.51 (IQR: 0.21-59.21) in those without parasites (p=0.764). In addition, 174/261 [66.7% (95% CI: 60.68-72.16)] and 231/328 [70.4% (95% CI: 65.23-75.14)] specimens from COVID-19 patients with parasite co-infection and without parasites, respectively, had anti-SARS-CoV-2 antibody above the cut-off COI value (p=0.328). The positivity rate for anti-SARS-CoV-2 NP < 14 days after symptom onset was 66.3% (95% CI: 60.21-71.85) and 70.0% (95% CI: 64.72-74.74) for those not infected and co-infected with parasites, respectively (p=0.343). In addition, 31/72 (41.9%) of the patients who were negative at the time of enrollment were seroconverted. The trend in anti-NP antibodies among seroconverted individuals with and without parasites is also similar.

**Conclusions:** Co-infection with parasitic infection has very little effect on the anti-SARS-CoV-2 antibody immune response. Further studies on the profile of neutralizing antibodies in parasite-endemic areas are warranted to ascertain vaccine efficacy.

**Author’s summary:** Pre-existing co-infection with intestinal parasites has been shown to diminish antibody response to a multitude of heterologous pathogens or vaccines. However, the effect of parasite co-infection on antibody response to SARS-CoV-2 is unknown. We determined the anti-nucleocapsid protein (NP) antibody response to SARS-CoV-2 among COVID-19 patients co-infected with intestinal parasites and compared their response to those without parasites. There was no difference in anti-NP positivity rate, seroconversion, or titer level among COVID-19 patients with or without parasitic co-infection. Further studies on the profile of neutralizing antibodies in parasite-endemic areas are warranted to ascertain vaccine efficacy.

## Introduction

Antibody testing for severe acute respiratory syndrome coronavirus-2 (SARS-CoV-2) is used for epidemiological surveillance, in aiding the diagnosis of (previous) coronavirus disease 2019 (COVID-19), and in the assessment of herd immunity following natural infection or after vaccine administration [1-9]. Several reports have demonstrated a lack of specificity for SARS-CoV-2 antibodies in evaluations of samples both pre-pandemic as well as samples collected during the COVID-19 pandemic period [10-18]. Such samples derived from parasite-endemic areas may cross-react to some of the SARS-CoV-2 antigenic epitopes [19, 20]. On the other hand, antibody response, including neutralizing antibody titers may be influenced in individuals with profound immune suppression, such as HIV-1 [21], though others have not demonstrated such effects [22, 23].

B-cell hypo-responsiveness is associated with intestinal parasite co-infection in endemic areas [24]. Indeed, pre-existing co-infection with parasites has been shown to diminish antibody response to influenza, hepatitis B, pneumococcal, cholera, tetanus, and malarial vaccines [25-31].

Nonetheless, the effect of parasite co-infection on antibody response to SARS-CoV-2 among COVID-19 patients remains to be elucidated. In addition, seroconversion might also be reduced or delayed in SARS-CoV-2-infected patients with pre-existing parasite co-infection. Here, we determined whether pre-existing co-infection with parasites impacts SARS-CoV-2 antibody responses and seroconversion.

## Materials and Methods

### Study design, period, and setting

The study is part of the ongoing Profile-Cov study, a prospective observational cohort study in Ethiopia with rapid diagnostic profiling of SARS-CoV-2 in the context of persistent immune activation (Clinicaltrials.gov: NCT04473365). Consecutive patients with confirmed Real–Time polymerase chain reaction (RT-PCR) test results were enrolled. They were recruited from Mekelle University College of Health Sciences (Kuyha COVID-19 Isolation and Treatment Center), Mekelle City, Northern Ethiopia, prospectively between July 15 and October 28, 2020.

Individuals presenting to the Isolation and Treatment Center were screened for SARS-CoV-2 infection with a nasopharyngeal swab and an RT-PCR confirmed patients. Following the declaration by the WHO that COVID-19 has become a pandemic, the Ethiopian Ministry of Health implemented a mass screening of all travelers, people who had come in contact with COVID-19 patients, those from high-risk settings (e.g. healthcare workers), as well as those with symptoms and signs suggestive of SARS-CoV-2 infection [32]. All cases with a confirmed SARS-CoV-2 infection were admitted to dedicated COVID-19 Isolation and Treatment Centers. Patients were admitted irrespective of clinical severity status. Whenever patients progress to severe or critical COVID-19, they were admitted to the intensive care unit (ICU). None of the patients received SARS-CoV-2 vaccination as they were enrolled before the introduction of a vaccine to the country.

### Data and specimen collection

Sociodemographic, clinical data, and laboratory data were collected using standardized Case Record Forms (CRFs) adapted from the International Severe Acute Respiratory and Emerging Infection Consortium’s (ISARIC) CRFs for emerging severe acute respiratory infections [33]. All data were then entered onto electronic medical records.

For analysis of anti-NP antibody kinetics, samples were drawn serially from 72 SARS-CoV-2 RT-PCR-confirmed patients. Overall, patients were followed for a median of 31 [interquartile range (IQR): 17-32] days, and ranging between 7 to 37 days. Whenever possible, samples were drawn every 3 days during follow-up. Thus, multiple samples were drawn per individual at a given time interval (0–3, 4–6, 7–9, 10–12, 13–15, 16–18, 19–21, 22–24, 25–27, 28–30, 31–33, 34–36, 37–39, and ≥ 40 days after onset of symptoms). Notably, only one sample per time interval was included from a single participant. Specimens were immediately transported to the Central Laboratory of the university hospital, and separated plasma was frozen at –80°C until further analysis.

### Laboratory assays

SARS-CoV-2 infection was confirmed by RT-PCR on samples obtained from nasopharyngeal swabs, as described previously [32]. The antibody profile in this study was determined using the Roche Elecsys electrochemiluminescent immunoassay (ECLIA) that targets the nucleocapsid protein (NP). Testing, including the cut-off index (COI) for anti-NP total immunoglobulin (Ig) titer was done following the manufacturer’s instructions, as described previously [34]. Seroconversion was ascertained if a patient with a negative antibody test at the time of enrollment becomes positive with the assays tested during follow-up. For estimating the day of seroconversion, the day’s post-symptom onset was determined for symptomatic cases. For asymptomatic patients, we added 6 days (median time between symptom onset and date of positive PCR testing among symptomatic cases) to the PCR testing date. Fresh stool sample specimens were also obtained for examination for parasites and ova. The analysis included direct microscopic examination, modified Ritchie concentration method, and Kato-Katz method, as described previously [35].

### Statistical Analysis

Baseline characteristics for continuous variables were expressed as the median with IQR, and for categorical variables as proportions. Categorical variables were compared using Fisher’s exact test or χ^2^ test, and continuous variables were compared by Mann–Whitney U or Kruskal–Wallis tests as appropriate. The primary outcomes for this study were anti-NP total Ig positivity rate or titers above the COI value, according to sampling time (days after symptom onset). The positivity rate was presented as proportion and 95% confidence interval (CI) limits. Data were analyzed using STATA (Statistical package v. 14.0, StataCorp, Texas, USA).

### Ethical considerations

Participants provided written informed consent to participate in the Profile-Cov study. The study protocol was reviewed and approved by the Health Research Ethics Review Committee (HRERC) of Mekelle University College of Health Sciences (#ERC 1769/2020). Written informed consent was obtained from all study participants. However, specimens used as controls from pre– pandemic period were specimens from biobank and individual study participant consent was not obtained. Nonetheless, all personal identifiers were–linked from the sources, and the HRERC has reviewed and approved it.

## Results

### General characteristics

Detailed demographic and clinical characteristics of the cohort have been described previously in detail in our published articles [34, 35]. For this sub-study, 72 PCR-confirmed COVID-19 patients were enrolled and contributed a total of 589 samples [median of 9 (IQR: 5 – 11) paired samples per patient] collected serially over time. Of the 72 COVID-19 patients, 39 (54.2%) were co-infected with intestinal parasites while 33 (45.8%) had no parasitic co-infection. Details in sociodemographic characteristics and prevalence of parasite co-infection are summarized in **Table 1**. Frequent symptoms were fever, dyspnoea, cough, and headache. Except for dyspnoea which was significantly higher among those without parasites, clinical symptoms were similar in both groups. In addition, the groups did not differ by age, sex, comorbidities, or severity.

**Table 1:** Clinical features among COVID-19 patients without, or with parasite co-infection.

| Characteristic | All patients<br>N=74 | Without parasite<br>N= 35 (47.3) | With parasite<br>N=39 (52.7) | p-value |
| --- | --- | --- | --- | --- |
| **Socio–demographic features:** |  |  |  |  |
| Male sex | 57 (77.0) | 26 (75.3) | 31 (79.5) | 0.595 |
| Age in years (median, IQR) | 35 (28-50) | 37 (28-55) | 34 (28-43) | 0.368 |
| **Clinical symptoms:** |  |  |  |  |
| Fever | 23 (31.1) | 12 (34.3) | 11 (28.2) | 0.573 |
| Dyspnea | 30 (40.5) | 19 (54.3) | 11 (28.2) | **0.023** |
| Cough | 34 (46.0) | 19 (54.3) | 15 (38.5) | 0.173 |
| Chest pain | 7 (9.5) | 3 (8.6) | 4 (10.3) | 0.805 |
| Sore throat | 5 (6.8) | 3 (8.6) | 2 (5.1) | 0.556 |
| Headache | 21 (28.4) | 13 (37.1) | 8 (20.5) | 0.113 |
| Nasal congestion | 5 (6.8) | 1 (2.9) | 4 (10.3) | 0.205 |
| Loss of smell and/or taste | 3 (4.1) | 1 (2.9) | 2 (5.1) | 0.621 |
| Nausea/vomiting | 5 (6.8) | 3 (8.6) | 2 (5.1) | 0.556 |
| Myalgia | 8 (10.8) | 4 (11.4) | 4 (10.3) | 0.871 |
| **Comorbidities** |  |  |  |  |
| **Comorbidity (at least 1)** | 21 (28.4) | 10 (28.6) | 11 (28.2) | 0.972 |
| Non–communicable diseases (NCDs) | 18 (24.3) | 8 (22.9) | 10 (25.6) | 0.780 |
| **Outcomes** |  |  |  |  |
| Severe COVID–19 | 29 (39.2) | 17 (48.6) | 12 (30.8) | 0.117 |
| Parasite co-infection | 39 (52.7) | - | - | - |
| Protozoa - any | 26 (35.1) | - | - | - |
| *Entamoeba spp. cyst* | 18 (24.3) | - | - | - |
| *Entamoeba histolytica trophozoite* | 5 (6.8) | - | - | - |
| *Giardia lamblia trophozoite* | 3 (4.1) | - | - | - |
| Helminth – any | 19 (25.7) | - | - | - |
| *Hymenolopis nana* | 15 (20.3) | - | - | - |
| *Schistosoma mansoni* | 2 (2.7) | - | - | - |
| *Ascaris lumbricoides* | 3 (4.1) | - | - | - |
| Poly–parasitism – any | 9 (12.2) | - | - | - |

\*Data are presented as proportion (%) unless indicated

### Longitudinal SARS-CoV-2 antibody response

The overall antibody NP total Ig response is summarized in **Table 2**. A total of 589 specimens derived at different time points were evaluated. The overall positivity rate for anti-SARS-CoV-2 total Ig was 68.8% (95% CI: 65.01-72.51); 70.4% (95% CI: 65.46-75.39) for those infected with parasites and 66.7% (95% CI: 60.91-72.42) for those not co-infected with parasites (p=0.328). Additionally, the positivity rate was 69.7% (95% CI: 62.61-76.78) for those infected with helminths and 68.4% (95% CI: 63.95-72.84) for those not co-infected with helminths (p=0.760). Whereas the anti-SARS-CoV-2 NP total Ig was 68.8% (95% CI: 65.01-72.51) among those infected with protozoa, it was 67.4% (95% CI: 62.62-72.13) among those without protozoa (p=0.333). Moreover, we did not observe differences in the anti-NP total Ig titers between those infected with parasites and those without parasite co-infection (**Table 2**).

**Table 2.** Profile of anti-SARS-CoV-2 NP total Ig response among COVID-19 patients with or without intestinal parasite co-infection.

| Anti-NP total Ig* | Parasite |  |  | Helminth |  |  | Protozoa |  |  |
| --- | --- | --- | --- | --- | --- | --- | --- | --- | --- |
|  | Negative | Positive | P-value | Negative | Positive | P-value | Negative | Positive | P-value |
| Positivity rate | 174/261<br>(66.7) | 231/328<br>(70.4) | 0.328 | 290/424<br>(68.4) | 115/165<br>(69.7) | 0.760 | 254/377<br>(67.4) | 151/212<br>(71.2) | 0.333 |
| Anti-NP total Ig titer<br>(COI) | 7.51<br>(0.21-59.21) | 6.91<br>(0.55-40.70) | 0.764 | 6.84<br>(0.28-48.99) | 7.49<br>(0.58-42.35) | 0.676 | 7.51<br>(0.23-50.57) | 6.79<br>(0.62-39.86) | 0.737 |

Data presented as numbers and percentage positivity rate, or median (IQR) cut-off index (COI). The COI for positive anti-NP total Ig is $\geq 1.0$ .

The kinetics of the anti-NP total Ig response are shown in **Figure 1**. The seropositivity rate was low at the time of enrollment in the study. Overall the positivity rate for anti-NP total Ig was 11.1% in those co-infected with parasites and 14.3% in those without parasites (**Figure 1A**). Similar trends were observed among those with or without helminth (**Figure 1B**), or with or without protozoa (**Figure 1C**). However, during the first 15 days after the onset of symptoms, the positivity rate increased significantly reaching above 50% positivity rate, irrespective of parasite co-infection status. During the period 2-to 3 weeks after the onset of symptoms, the positivity rate of all the assays was increased, ranging between 67.3% and 81.3%. Between 3 and 6 weeks after the onset of symptoms, the positivity rate continued to increase for all the assays, ranging from 84.6% to 90.9%. Positive anti-NP total Ig titer was detectable (i.e. COI ≥ 1.0) as early as 7 to 12 days after onset of symptoms for all patients (**Figure 1D–F**). Because of the small sample size, we did not determine trends stratified by severity status.

**Figure 1.**
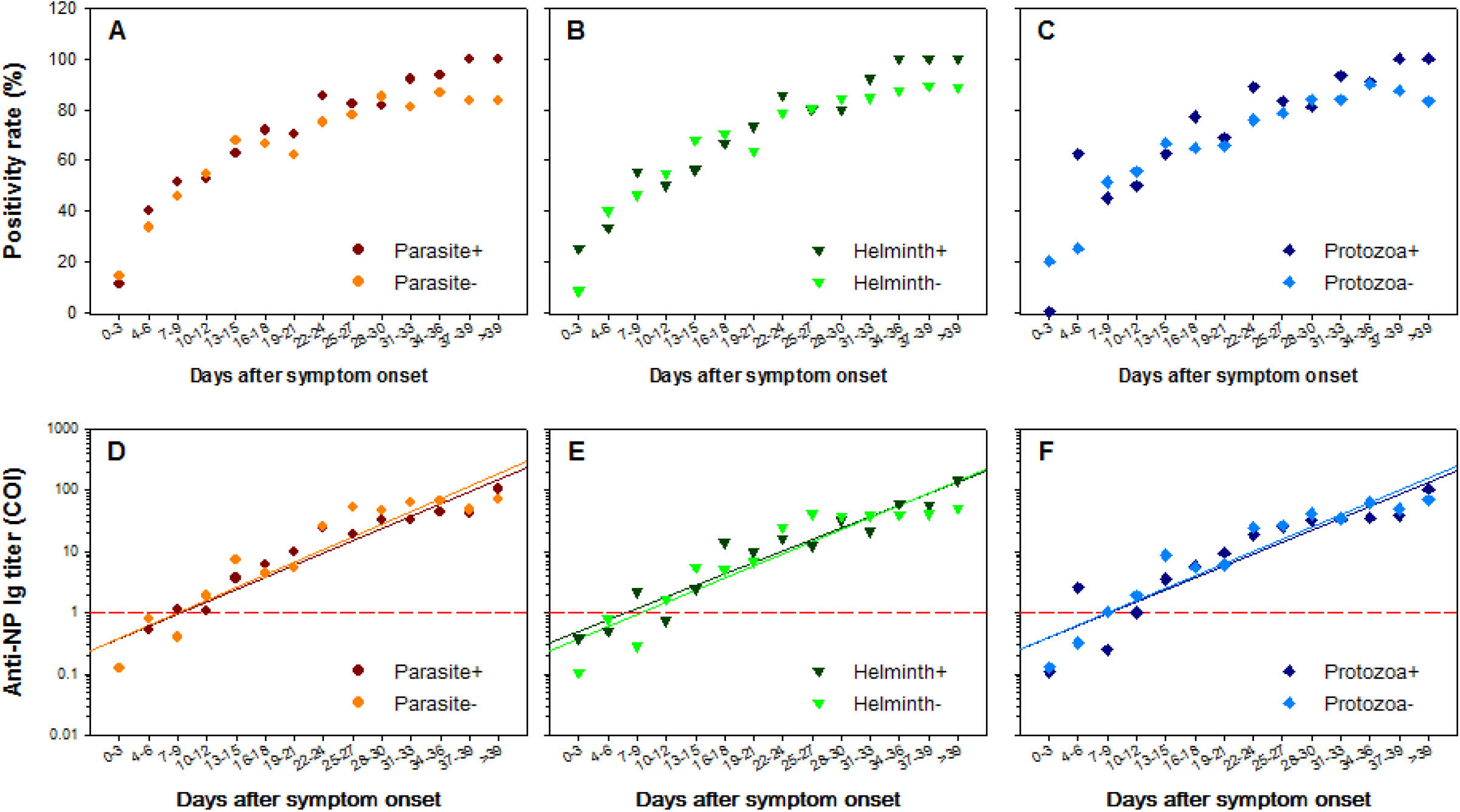
Anti-NP total Ig positivity rate (A-C) and titer levels (D-F) at different time points after onset of symptoms stratified by the parasite (A, D), helminth (B, E) and protozoa (C, F) co-infection status. The red dashed horizontal line (D-F) represents the cut-off value of the limit of detection of the anti-NP total Ig assay.

### Seroconversion of SARS-CoV-2 antibodies

Patients who were antibody–negative at the time of enrollment in the study, but who became antibody–positive during follow–up were considered seroconverters. Overall, a total of 31 (41.9%) were seroconverted. A total of 256 sample pairs were evaluated. Seroconversion trends are shown in **Figure 2**. Patients started to seroconvert as early as 4 to 9 days after the onset of symptoms, reaching around 25%. The rate of seroconversion within the first 15 days after the onset of symptoms increased to 44.4%. During the same period, the rate of seroconversion for those with parasites was 47.1% (95% CI: 20.61-76.94), and for those without parasitic co-infection was 40.0% (95% CI: 3.06-76.94), and the difference was not statistically significant (p=0.722). In addition, the seroconversion rate increased further during 2-to 3 weeks after the onset of symptoms, ranging between 61.5% and 68.0%, respectively. The seroconversion rate continued to increase reaching 100% after ≥ 39 days of symptom onset in all groups (**Figure 2A**). Trends in anti-NP total Ig titers were also similar in patients irrespective of the presence of parasite co-infection or not (**Figure 2B**). Whereas the median seroconversion time for those with parasites was 13 (IQR: 7-21) days, it was 15 (IQR: 10-16) days for those without parasitic co-infection (p=927). Overall, there was no difference in seroconversion trend between those who were co-infected with parasites and those without parasites. Due to the small sample size, however, we did not do an analysis stratified by helminths and protozoa.

**Figure 2.**
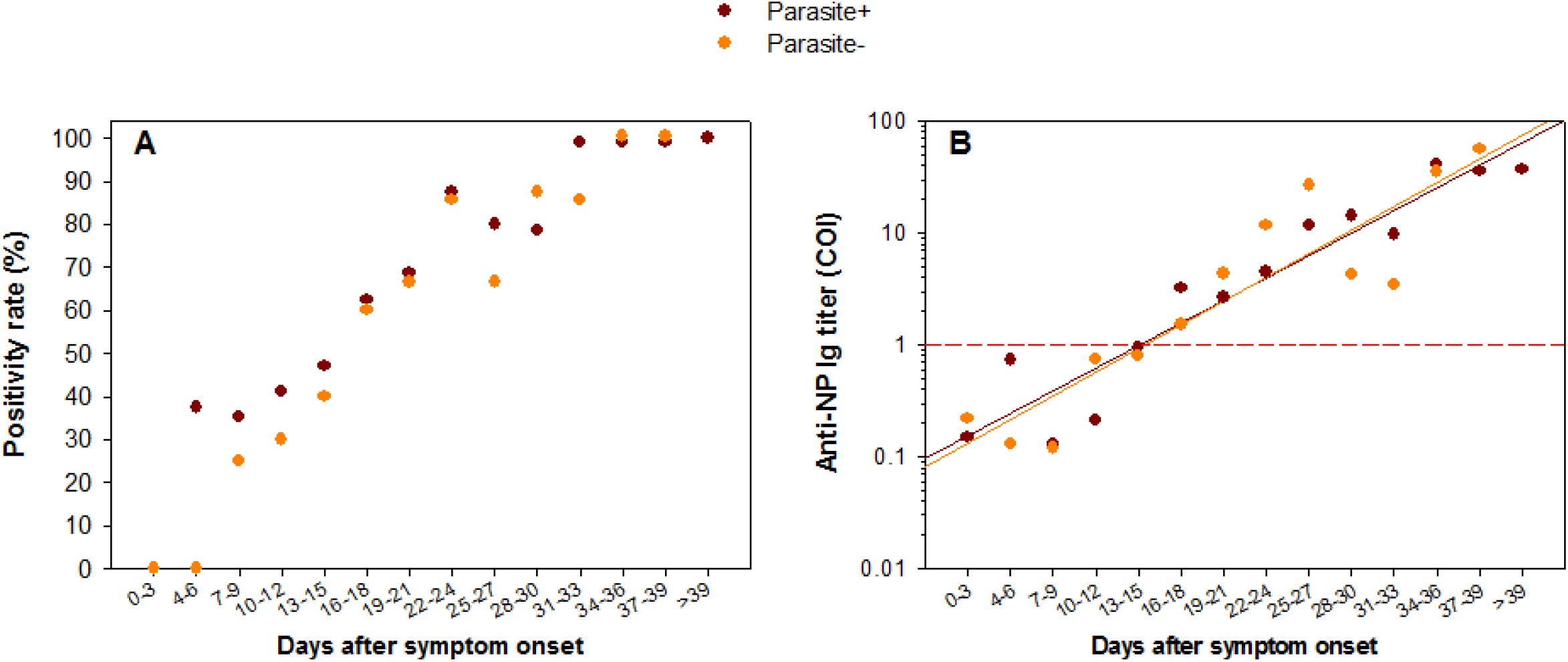
Seroconversion rate (A) and titers levels (B) of anti-NP total Ig at different time points after onset of symptoms stratified by parasite co-infection status. The red dashed horizontal line (B) represents the cut-off value of the limit of detection of the anti-NP total Ig assay.

## Discussion

In this study, we determined antibody responses among SARS-CoV-2 patients co-infected with parasites. To the best of our knowledge, this report is the first to determine the longitudinal SARS-CoV-2 antibody response in COVID-19 patients also co-infected with intestinal parasites. SARS-CoV-2 patients with parasite co-infection did not exhibit different profiles in antibody response or seroconversion patterns.

Several reports have previously documented that parasites downregulate antibody responses to heterologous pathogen and vaccine responses [25-31]. Likewise, we expected an impaired antibody response to SARS-CoV-2 among COVID-19 patients co-infected with parasites [24]. Given the presence of a high level of cross-reactivity between SARS-CoV-2 and parasites [10-20], the expected reduction in antibody response might have been compensated by the presence of cross-reacting antibodies also in patients with COVID-19. Furthermore, it has been demonstrated that a reduced frequency of SARS-CoV-2-reactive CD4+ T helper cells without an effect on the expression of SARS-CoV-2-reactive CD8+ T cells [36]. Thus, it is plausible to postulate that parasite-induced reduced CD4+ T helper cells do not affect anti-NP antibody responses to SARS-CoV-2.

The strength of the current study is the use of longitudinal samples derived from the same patient. However, limitations include smaller sample sizes and a lack of determination of neutralizing antibody titers. In addition, anti-NP antibody response has not been stratified by severity status, though in our set-up we did not find differences in antibody profile by severity status [34].

The impact of parasite co-infection on the efficacy of SARS-CoV-2 vaccines in endemic areas is unknown [37]. Unraveling the differential antibody response profile between individuals with the parasite and those without parasite co-infection may have implications in the development and interpretation of diagnostic tools as well as the monitoring of vaccines in parasite-endemic areas. Further studies are needed to investigate the effects of co-infection with parasites on the efficacy and durability of antibody responses to SARS-CoV-2 vaccines.

## Data Availability

All relevant data are within the manuscript and its Supporting Information files.

## Informed Consent Statement

Written informed consent was obtained from all study participants. All personal identifiers were de–linked from the sources.

## Acknowledgments

The investigators thank EDCTP for providing funding to the project, COVID-19 patients and all individuals who provided specimens for control, and the medical staff who provided care of the patients.

